# Globally Local: Hyper-local Modeling for Accurate Forecast of COVID-19

**DOI:** 10.1101/2020.11.16.20232686

**Authors:** Vishrawas Gopalakrishnan, Sayali Pethe, Sarah Kefayati, Raman Srinivasan, Paul Hake, Ajay Deshpande, Xuan Liu, Etter Hoang, Marbelly Davila, Simone Bianco, James H. Kaufman

## Abstract

Multiple efforts to model the epidemiology of SARS-CoV-2 have recently been launched in support of public health response at the national, state, and county levels. While the pandemic is global, the dynamics of this infectious disease varies with geography, local policies, and local variations in demographics. An underlying assumption of most infectious disease compartment modeling is that of a well mixed population at the resolution of the areas being modeled. The implicit need to model at fine spatial resolution is impeded by the quality of ground truth data for fine scale administrative subdivisions. To understand the trade-offs and benefits of such modeling as a function of scale, we compare the predictive performance of a SARS-CoV-2 modeling at the county, county cluster, and state level for the entire United States. Our results demonstrate that accurate prediction at the county level requires hyper-local modeling with county resolution. State level modeling does not accurately predict community spread in smaller sub-regions because state populations are not well mixed, resulting in large prediction errors. As an important use case, leveraging high resolution modeling with public health data and admissions data from Hillsborough County Florida, we performed weekly forecasts of both hospital admission and ICU bed demand for the county. The repeated forecasts between March and August 2020 were used to develop accurate resource allocation plans for Tampa General Hospital.

**2010 MSC:** 92-D30, 91-C20

## Introduction

The COVID-19 pandemic caused by the SARS-CoV-2 virus reveals the important contributions that can be made by accurate mathematical modeling of infectious disease. A variety of modeling approaches can be used to predict the course of an infection in a population. While the pandemic is global, the underlying disease dynamics (and the epidemiological parameters themselves) varies with specific local policies, non-pharmaceutical interventions (NPI), variations in demographics, healthcare infrastructure, etc. To realize the full potential of any modeling effort, forecasts and predictions must be accurate and meaningful to public health officials and governing authorities at a resolution relevant to their areas of responsibility^1^. In most cases, this requires predictions be made at a hyper-local level. Models can only be used to guide closing or opening of business, predict and allocate Intensive Care Units (ICU) resources, and establish social distancing guidance if their predictions are meaningful at a scale relevant to these businesses, hospitals, and communities.

In practice, the ability to model at high resolution is limited by the granularity and accuracy of ground truth data, as well as the underlying modeling assumptions^2,3^. Precision of public health SARS-CoV-2 case reports, including incidence and death^4^, is impeded by multiple noise sources^5^. Clinical encounters may take place adjacent to or outside a patients residential district, testing capacity, public health reporting practice, and many other factors may vary dramatically even between nearby districts. Some of these noise sources may be reduced by clustering administrative divisions or modeling at lower resolution, but this approach may violate an assumption used by most compartmental models, that the population being modeled is well mixed^6^.

In this paper, we compare the predictive performance of a SARS-CoV-2 model as a function of spatial scale, applying the model to county, county cluster, and state level divisions. For this purpose, we selected an epidemiological compartment model, as opposed to a regression model, to investigate changes in important disease parameters obtained at each resolution, and to avoid an approach where important parameters are under determined. Predictive power and scaling behavior are expected to depend on model selection. An open source model and modeling framework were chosen so others can reproduce the results and test future extensions that may improve model performance. To evaluate the performance of the selected model we also compare the accuracy to other models with predictions publicly available through the US Center for Disease Control and Prevention (CDC)^7,8^. These other models include both epidemiological compartment models, regression models, and hybrid approaches. Our results reveal that, for the United States, even for fairly small states, different counties make dominant contributions to daily incidence at different times. The mean average percentage error (MAPE) in predicted incidence can be 30% higher for modeling at the state level compared to aggregating model results at the scale of counties or clusters of counties. Furthermore, county level predictions made by partitioning state level predictions (for a model performing well at the state level) are 3x worse for predicted incidence and 20x worse for predicted deaths relative to the same model at the county level. These results demonstrate that modeling resolution must be guided by the requirements of the consumer of the predictions. State level modeling strongly violates the assumption of well mixed populations and will lead to inaccurate predictions at a county level even while performing well at the state level. In order to test the performance of the COVID-19 models at the county level, IBM partnered with Tampa General Hospital (TGH) to forecast COVID-19 incidences within Hillsborough County. Hillsborough County accounts for a large portion of the Tampa Florida population. Tampa General Hospital (TGH) is the only Level 1 Trauma Center within the Tampa region. As COVID cases rose in early March, TGH has treated an average of 20.4 percent of the Hillsborough COVID-19 cases with a max distribution of 45.0 percent of all COVID-19 cases in Hillsborough County. The forecasting models would become imperative to ensure adequate strategic planning for hospital capacity, staffing, and supplies as the virus spread.

## Methods

### Data Description

USAFacts COVID-19 datasets^9^ including daily COVID-19 incidence and deaths, were obtained directly from CDC, state and local agencies and used for model optimization. This data is further curated; for instance, the post-processing ensures that the number of confirmed cases is a monotonically increasing curve. The data is refreshed every day around 9AM PST to reflect updates of the previous day. More information on the reference data^10,11,12^, and on the CDC challenge models^7^ used for model comparison is available in the supplement.

### Base Model

For this study we chose a modeling framework, the SpatioTemporal Epidemiological Modeler (STEM)^13,14^, available through the Eclipse Foundation^15^. The framework and model are open source and available under the Eclipse Public License (EPL2)^16^. A number of models for SARS-CoV-2 from multiple authors were evaluated^17,18,19^. For this study we use the model shown in figure 1, which includes transmission from both asymptomatic (*A*), pre-symptomatic (*C*), and infectious individuals (*I*). Moreover, we capture infectious individuals who experience a worsening of symptoms (*W*) before either dying of the disease (*D*) or recovering (*R*). This model is a simple extension to the SACIR model available on the Eclipse site.

**Figure 1:**
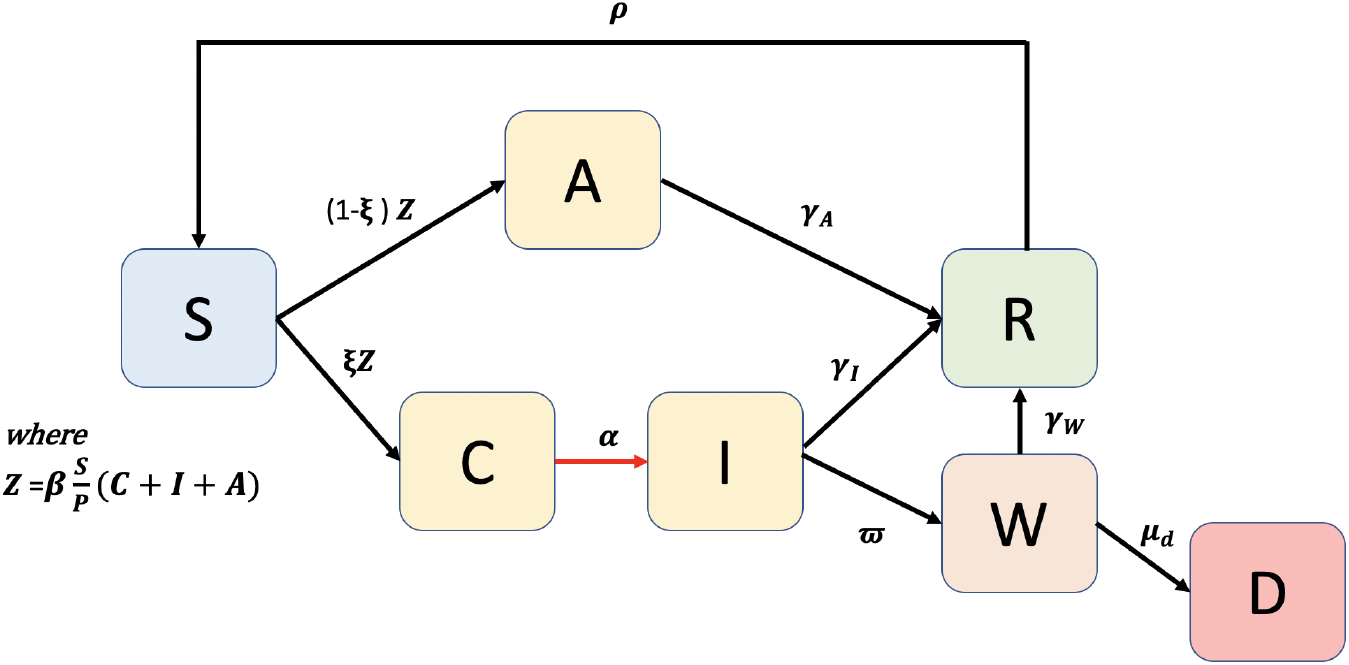
Compartment model use to capture the disease dynamics of SARS-CoV-2 (see text).

Model parameters and initial conditions were selected by a combination of literature references, grid search, and data-driven estimation through a Nelder-Mead AI simplex algorithm available in the STEM framework. The flow diagram of figure 1 translates into a system of ODE’s reported in the supplemental Eq. 2. The definition of each compartment, all model parameters, literature values, and methods of parameter estimation are discussed in the supplement. The basic epidemiological model was used to forecast at three spatial resolutions (counties, county clusters, and states). Definition of county clusters is described in the supplement.

Model performance was measured using the normalized root mean square error (nrmse) of daily incidence, cumulative incidence, and cumulative deaths with respect to the corresponding ground truth data one to four weeks into the future. Since public health reporting usually takes place after a clinical encounter, the transition between the pre-symptomatic compartment (*C*) and the infectious compartment (*I*) was used to log daily incidence (red arrow, figure 1). Here we introduce a pre-symptomatic compartment *C* in place of the usual exposed compartment *E* based on documented evidence for replication-competent virus obtained from patients before the onset of symptoms^20,21,22,23,24^. Thus, we describe the rate of individuals leaving the *C* compartment as a symptom appearance rate as opposed to an incubation rate. The processes are similar in that they both would contribute to a period of latency between exposure and clinical encounter. We chose the nomenclature to indicate that individuals in the *C* compartment contribute to the disease force of infection.

The steady state solution to the differential equations 2 shown in the supplement provides the following expression for the basic reproductive number, *R*_0_ as a function of the epidemiological parameters. Background birth and death rates are omitted. Individuals in the W compartment do not contribute to the force of infection.

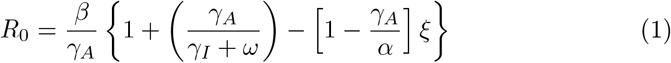

To provide a scalable architecture for modeling, we built an automation pipeline that performs a variety of tasks ranging from data ingestion, smoothing, model invocation, and post-processing. The pipeline supports multi-processing, where each region is run by a separate instance of a docker image; allowing easy scaling of the clusters as required. This pipeline is discussed in the supplement.

### Hospitalization and Intensive Care Unit Forecasting Model

In order to deliver actionable projections for decision making by healthcare providers, as a post process we decomposed the incidence projections using rate equations to forecast both total hospitalization and ICU demand at the county level. The average rate (weighted by incidence) of hospitalization per case (incidence) was 0.35±0.04 and the average rate of ICU admission per hospitalized patient as 0.10±0.05.

Tampa General Hospital in Hillsborough County, Florida, USA, implemented this model to anticipate future resource demand from COVID-19 patients and to advise decisions regarding possible need for extra capacity and reduction in planned surgeries to free up bed space. They established a regional collaboration for data and resource sharing facilitated by shared data collection and a reporting dashboard. More information on this model is available in the supplement. Projections based on both models were updated at least weekly since April 2020.

## Results

### Derived Epidemiological Parameters

The model optimization process automatically adds discrete changes to the transmission rate *β*, at discrete times, based on the dynamics observed in ground truth incidence and deaths. In an attempt to distinguish between a “natural” range of transmission rates, *β*, reflecting human contact rates in the absence of interventions, and the larger range of *β* with interventions, the violin plots in figure 2a are split. The left hand (cyan) distribution includes only the maximum *β* observed by region. The right (purple) distributions include all *β* values (i.e. all changes in *β* from local policies and NPI, as well as the maximum values). Similarly the distribution of *R*_0_ derived from equation 1) is shown in figure 2b. These distributions are also split with the left hand violin displaying *R*_0_ values computed from maximum *β* values observed by region. The right hand violin plot (labeled *R*_*eff*_) includes reproductive numbers computed independently from *all β* values observed in all regions. The distribution of transmission rates and reproduction numbers are multi-modal reflecting differences in disease dynamics between regions. Distributions for other parameters and a table listing L-statistics for all optimized parameters and reproduction numbers may be found in the supplementary Table 4 and 5 respectively. The Table also shows the average model convergence error (nrmse) which was under 4% for simulation at the scale of U.S. States, systematically increasing to just over 10% for simulation of U.S. counties.

**Figure 2:**
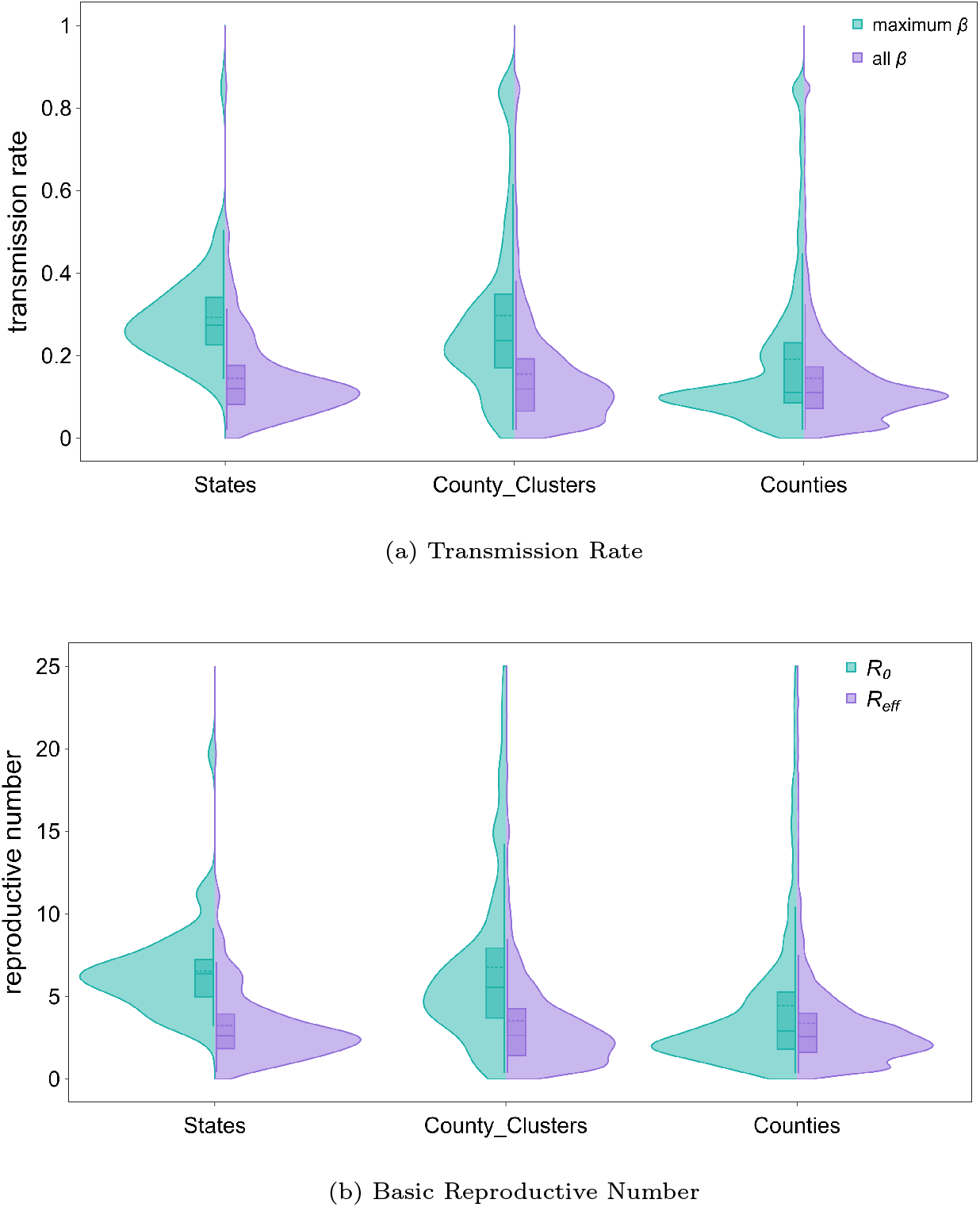
Regional distributions of transmission rates and reproduction numbers at different modeling resolutions. The violin plots are split to visualize both the distribution of maximal regional values and all regional values to show how non-pharmaceutical interventions may have lowered transmission rates and reproductive numbers (see text).

The optimized parameter values and derived values for the basic and effective reproduction numbers are within the range estimated from current clinical studies.^20,21,22,23,24^ Supplementary Table 5 provides statistics from the measured distribution of reproduction number values obtained using the epidemiological parameters in Table 4 at each spatial resolution.

### Model Performance and Predictions

Figure 3 shows example output of the epidemiological model for two states. The model was run at county, county cluster, and state resolution for all states (and DC). Figure 3 shows the actual and predicted (daily) incidence and (cumulative) deaths vs. time all county clusters within each state respectively. The data is presented as a stack chart so the envelop over aggregated clusters reflects total incidence and deaths at the state level. This is also indicated by the blue line above the stacked sub-regions for both the actual (reported) incidence and deaths, and the corresponding predictions. In each sub-figure the white background indicates the reference time period and data used for model optimization. The gray background shows actual and predicted incidence and deaths for subsequent days. This latter region was used to compute the MAPE.

**Figure 3:**
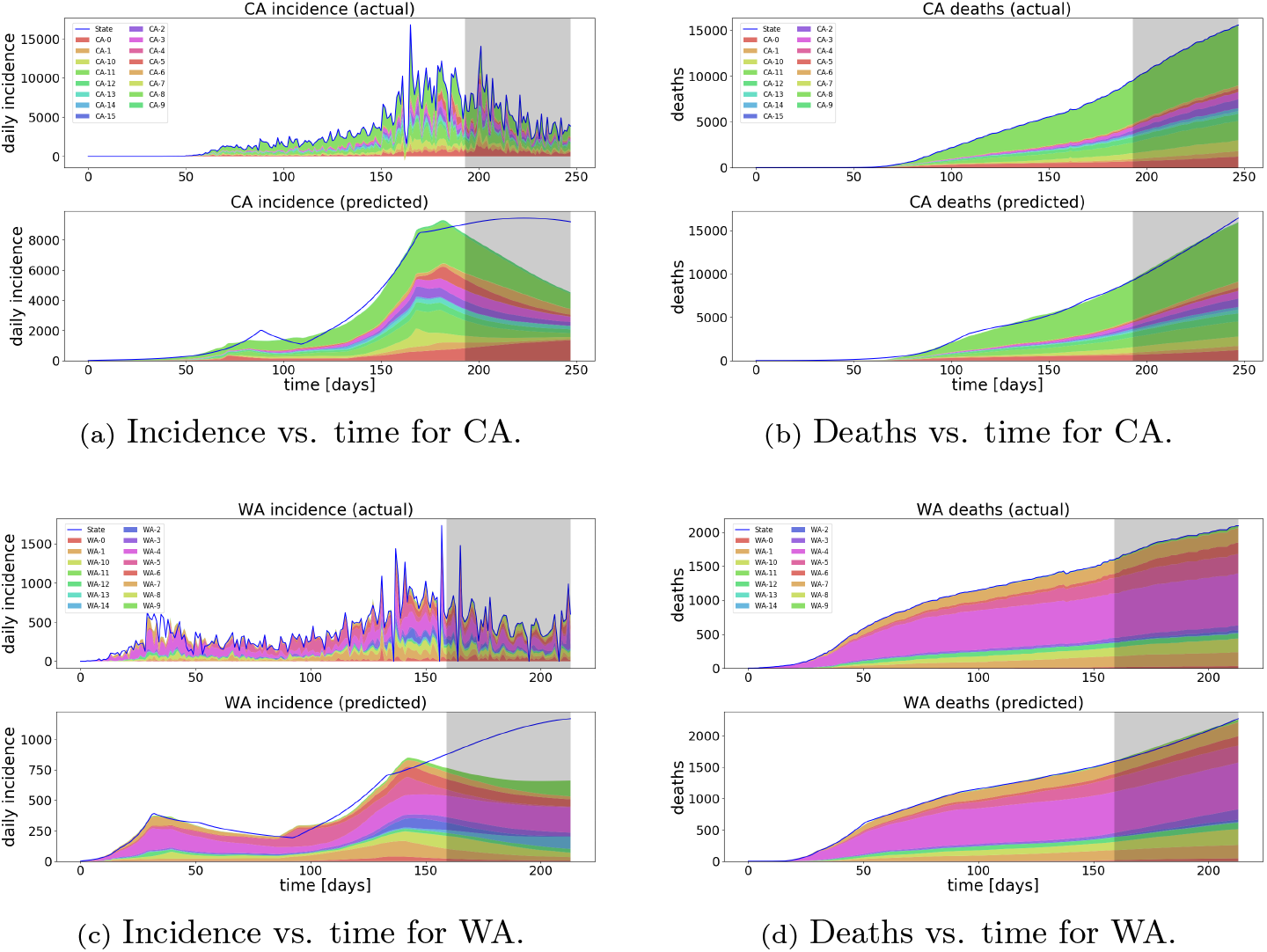
Actual and predicted (daily) incidence and (cumulative) deaths vs. time for county clusters within states. The white background shows the time period used for model optimization. The gray background shows incidence and deaths for subsequent days. The relative contribution to state level incidence or deaths from different sub-regions varies over time.

The data for California and Washington State (figures 3a and 3c) illustrate how the noise in daily incidence can adversely impact predictions made by an automated pipeline. This is evident as the model predictions at the state level (blue curve) deviate from the ground truth data after a particularly large noise spike. Predictions at individual and state aggregate county clusters do not show the same deviation. The stack charts also reveal that the relative contribution from different sub-regions to state level incidence or death varies over time indicating the states themselves are not accurately described as well mixed.

Prediction accuracy as a function of spatial scale is expected to be model dependent. Therefore, before evaluating the effect of modeling resolution on statistical error, we sought to test the predictive accuracy of the SACIR model relative to other models at the same resolution to establish its baseline performance. For this purpose we selected models from the CDC challenge project^7^. The majority of these models provide predictions at the state level. The pipeline predicts daily and cumulative incidence and cumulative deaths for all states at least four weeks into the future. Figure 4a, shows a self-consistent comparison of the accuracy of prediction, at state level resolution, for all models that predict both incidence and deaths at state level, for all states, and with data provided 1-4 weeks into the future. As discussed in the Methods section, some models were omitted from the comparison based on missing data.

**Figure 4:**
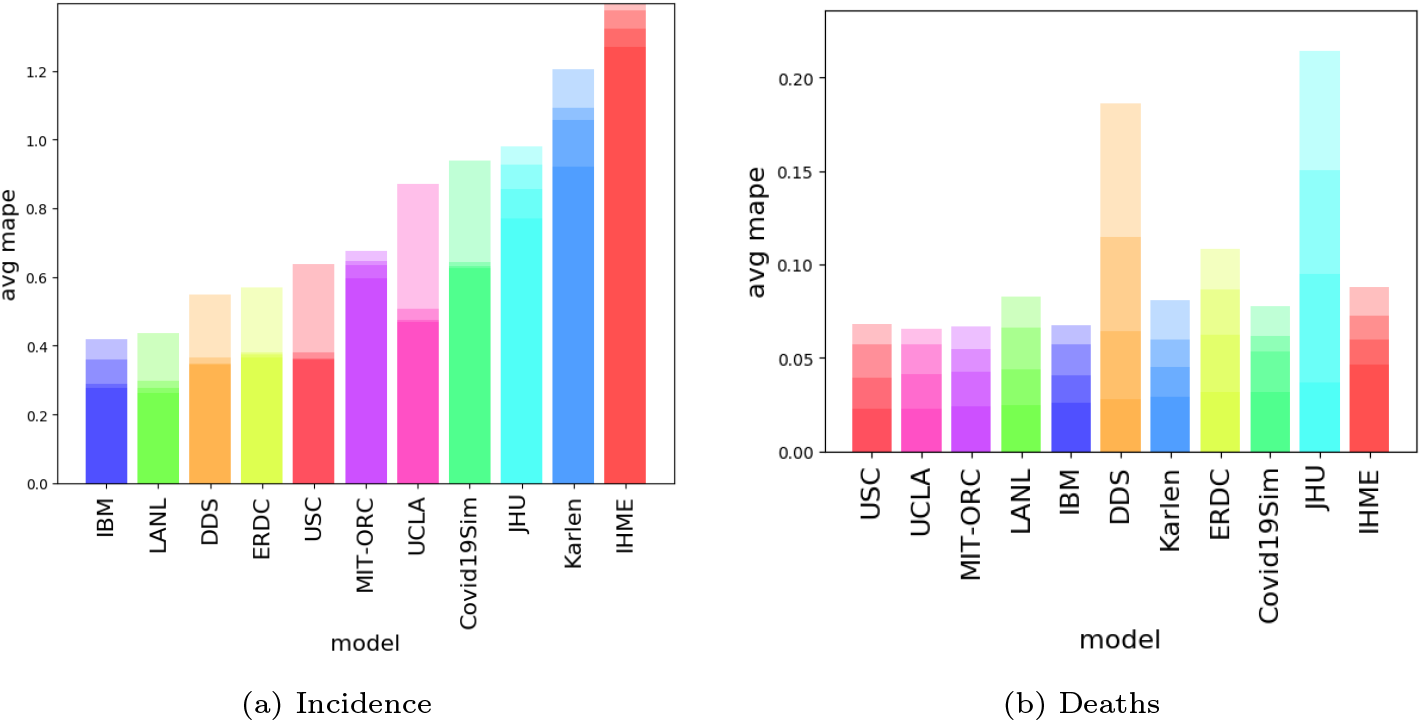
Mean absolute percentage error (MAPE) by model after 1-4 weeks. Models are sorted by prediction accuracy after one week

Figures 4a and 4b compare the statistical accuracy of several models, averaged over all states, at one to four weeks beyond the time of optimization. The order of each bar is based on the MAPE (low to high) after the first week. For each model the average MAPE increases with time after the optimization window. The baseline predicted performance, when run a state level resolution, of the SACIR model compares favorably with the other CDC Challenge models run at the same resolution for the same ground truth data and time.

### Effects of Spatial Scale

Given the state level baseline from figure 4a, we next compare the MAPE for predictions as a function of spatial scale. Figures 5a and 5c test the assumption that spatial regions defined by states represent well mixed populations. The figures show the statistical error in predicted incidence and predicted deaths, respectively, as a function of time. The red curves correspond to the average MAPE for models run at the state level (i.e. treating each state as single well mixed regions). The blue curves correspond to the average over states of the state level MAPE for models run at the county-cluster level. In this case the prediction for each state was obtained by summing all of the county cluster level predictions within the state, and the state level MAPE computed by comparing the sum to the ground truth data for the state. The green curves were computed in the same way as the blue curves but running the model at county resolution. From figure 5a it is clear that modeling at either county or county-cluster resolution lowers the mean absolute percent predictive error by 30% for each of the four future weeks. County and county cluster resolutions yield comparable results. All three resolutions provide comparable predictive accuracy for cumulative deaths (figure 5c) at least in the first two weeks. State level prediction accuracy of cumulative deaths degrades slightly faster after two week.

**Figure 5:**
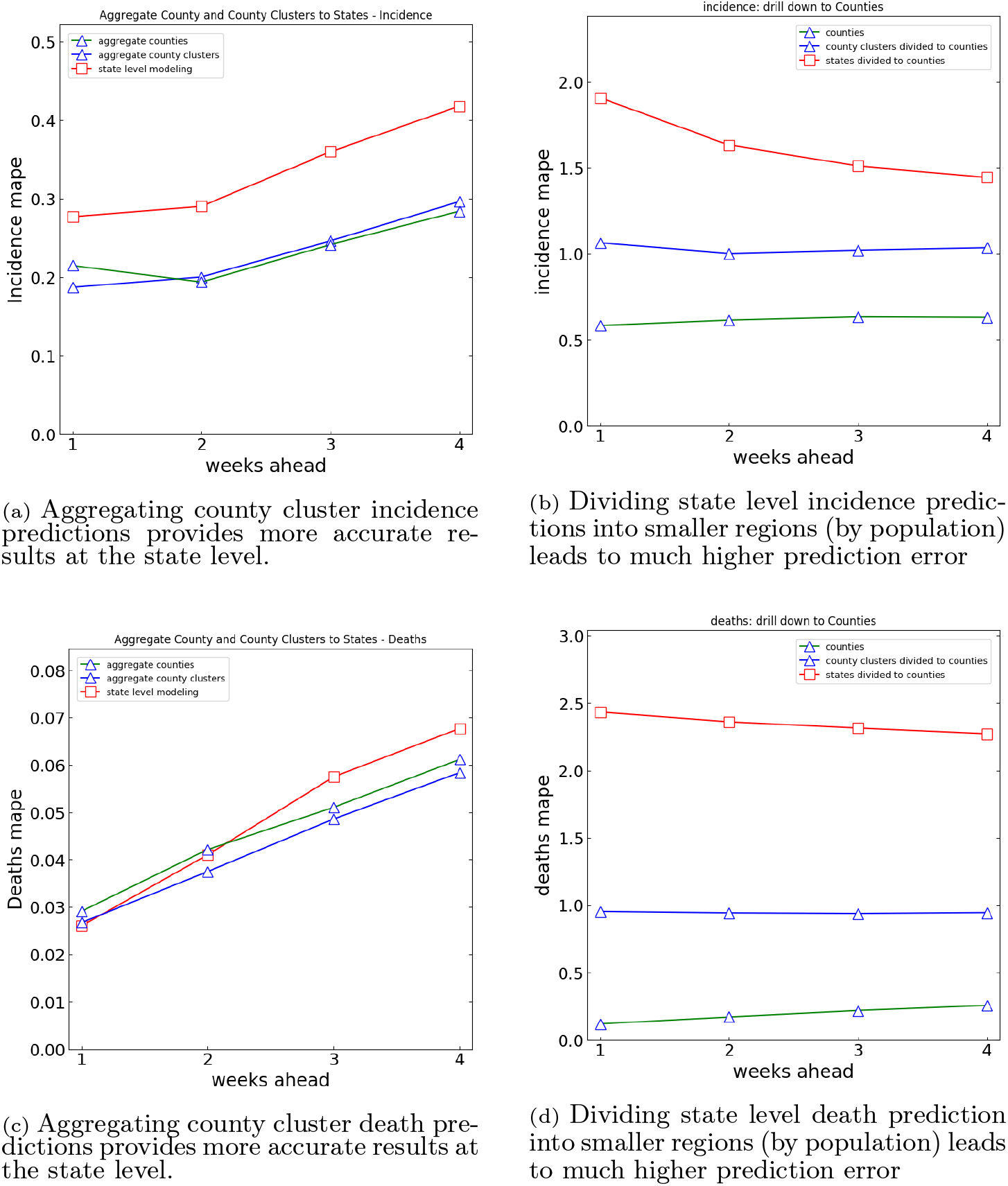
Comparing aggregation of smaller counties or county clusters provides more accurate state level prediction of deaths and incidence. Conversely, subdividing state level model results into counties or county clusters fails, demonstrating that most states are not approximated as well mixed regions.

Figures 5b and 5d test the complementary hypothesis. If larger regions are truly well mixed, then prediction at at higher resolution could be obtained by simply dividing or distributing predictions made for larger regions by the fraction of population in each sub-region. Figures 5b and 5d show the average MAPE for three methods to predict incidence or deaths at the county level. In each of the two figures, the red curve shows the average MAPE for county level predictions obtained by dividing the state level model output to obtain county level predictions. The blue curve represents county level predictions obtained by dividing county cluster level model output. The green curve is simply the average MAPE from modeling at the county level itself (not aggregated by state as in figures 5a and 5c).

From the data in figures 5b and 5d, we list in Table 1 the ratio of MAPE in prediction of county incidence or deaths based on state or county-cluster regions relative to modeling at the county level. The error ratios decrease with time (prediction week) as the MAPE increases for all resolutions over time. Looking at predictions one week into the future, county level predictions based on state level modeling have *∼* 3.3x higher error for predicted incidence and over 20x higher higher error for predicted deaths. Modeling counties at the county-cluster level, compared to county level, results in 1.8x higher error for predicted incidence and over 8x higher higher error for predicted deaths.

**Table 1:** Table increasing MAPE (ratio) as spatial resolution is decreases from county to state (see Figures 5b and 5d.

| Region Resolution | Week 1 | Week 2 | Week 3 | Week 4 |
| --- | --- | --- | --- | --- |
| Incidence |  |  |  |  |
| state/county | 3.3 | 2.7 | 2.4 | 2.3 |
| county-clusters/county | 1.8 | 1.6 | 1.6 | 1.6 |
| deaths |  |  |  |  |
| state/county | 20.6 | 14.0 | 10.6 | 8.9 |
| county-clusters/county | 8.1 | 5.6 | 4.3 | 3.7 |

To demonstrate the practical implications of our analysis, we report the fore-casts created to predict hospital admissions, and ICU demand for the Tampa General Hospital (TGH) network. Forecasts used predicted incidence data for Hillsborough County, and were performed at least weekly beginning in April 2020. Figure 6 shows actual and predicted COVID-19 ICU patients (magenta and blue solid curves respectively). The blue points indicate independent future projections. Total ICU patients, actual and predicted, are indicated by magenta and blue dashed curves respectively. Total ICU prediction in figure 6 was made by adding the predicted COVID-19 demand to a seven day moving average of non-COVID actual cases. The blue zone above and below the total ICU prediction represents the MAPE. The MAPE over the entire time range was 5.9%. The time-varying ICU bed capacity for the county is shown in red. Predicted and actual total ICU demand reached a maximum in June (before the peak COVID-19 demand) and never exceeded the ICU bed capacity of *∼* 350 beds at the time. Based on the total ICU demand projections, the hospital made the decision not to cancel other elective procedures and surgeries in the time period shown. Projections for county incidence, total hospital admissions, and statistical error (MAPE) for individual projections are available in the supplement.

**Figure 6:**
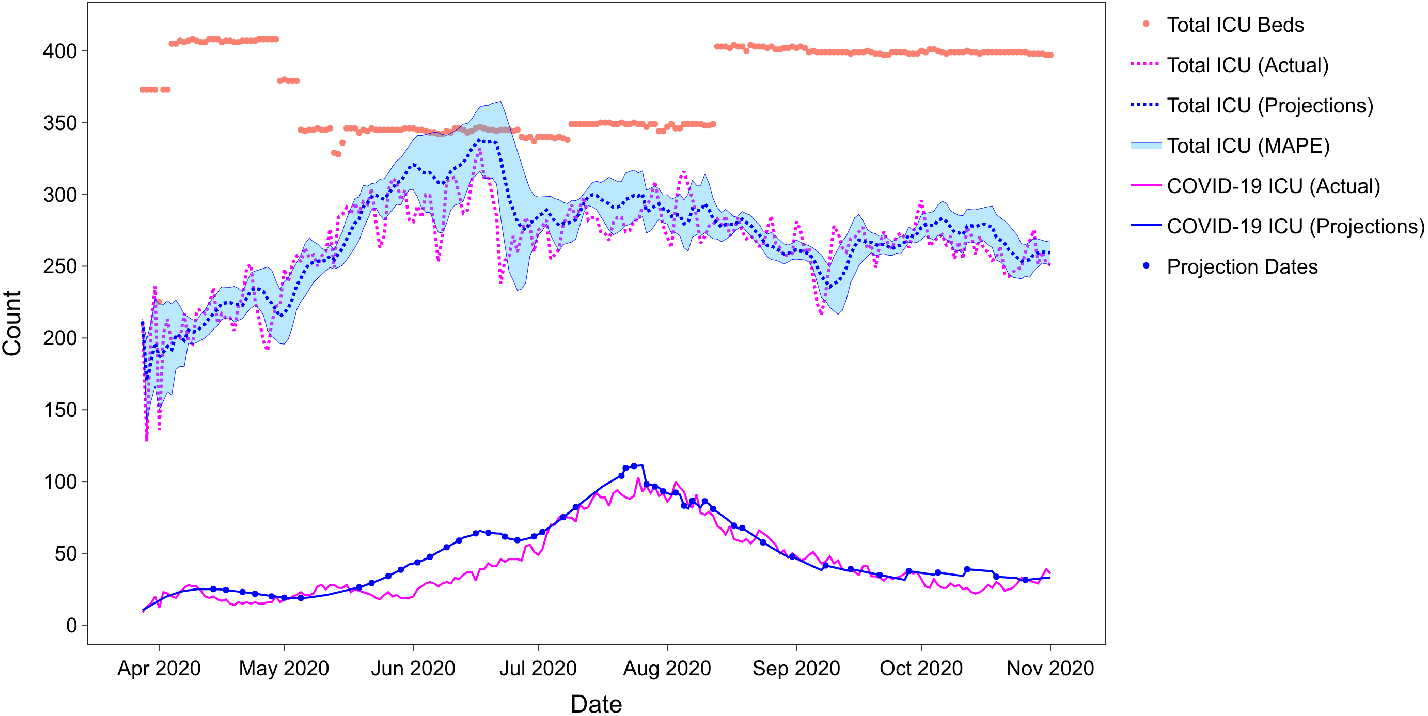
ICU Status for Hillsborough County, FL. The red points indicated capacity (which varied over time). The figure shows COVID-19 ICU Patients, all ICU Patients, and the full series of COVID-19 ICU projections made between April and November 2020.

## Discussion

The data in figures 4a and 4b provides a quantitative comparison of the performance of several different models, including the current SACIR model, based on the mean absolute percent prediction error one to four weeks into the future. These models each use different approaches and make different assumptions. Some are compartmental epidemiological models, some are based on regression, and some adopt a hybrid approach. The goal of this paper is not to identify a best model or paradigm, but to explore the question of modeling resolution itself.

Figures 4a and 4b compare the predictive power of models at several spatial resolutions. Modeling at a spatial scale corresponding to administrative divisions of states may have advantages in terms of reduced computational complexity and cost. While public health reports by county, lowering spatial resolution allows aggregation of inherently noisy data which contributes to computational complexity and cost. However, if public health officials and policy makers require accurate insights at county level, modeling at the state level may not suffice.

Figure 3 and associated data in the supplement demonstrates - from raw data alone - that different sub-regions within states make dominant contributions to aggregate state level incidence and deaths at different times. Figure 5 and Table 1 show that the statistical error (MAPE) in predicted incidence at the state level is 30% higher for modeling states compared to modeling at the level or counties or clusters of counties. Furthermore, testing the well mixed population assumption, the same figures reveal that county level predictions made by partitioning state level predictions (for a model performing well at the state level) are 3x worse for predicted incidence and 20x worse for predicted deaths relative to the same model at the county level. Figures 5a and 5c do indicate the possibility of diminishing returns as average MAPE in predicted incidence and deaths has similar (or even slightly lower) error at county-cluster resolution than at county resolution. Aggregating data to county clusters does provide noise reduction. Some of this noise is associated with artifacts of public health case reporting where incidence may be reported based on clinical location as opposed to patient residence. In other cases reporting location may be skewed by local public health practices. This is evidenced by public health data made available early in the pandemic where reported case counts and deaths were zero in Bronx, NY, while the epidemic grew exponentially in all adjacent boroughs^25^. Local reporting bias suggests that the optimal trade-off between noise reduction and spatial resolution may vary locally and even change over time as public health systems become more efficient over the course of an extended pandemic. Whatever decision one makes in optimizing modeling performance, the measure of success must be based on the accuracy achieved at the spatial resolution required by the consumers of the data. In the case of TGH Hillsborough, modeling at the county level the statistical error was < 6% (figure 6). Had the county prediction been based on state level modeling, the uncertainty would have increased by a factor of *∼* 3.3x to almost 20% (see Table 1), greatly reducing confidence in the forecast.

With relative accuracy from the model performance, TGH was able to consume the COVID forecasting model in two primary ways. In mid-June, as COVID-19 cases began a rapid rise in Hillsborough County, TGH leadership faced a difficult decision on whether or not to begin canceling operations to make capacity for the surging COVID-19 cases. As demonstrated in figure 6, the model accuracy improved in June between actual vs forecast, and predicted a peak incidence on 7/20/2020 and peak ICU on 7/26/2020. TGH then used the Hillsborough COVID model projections to define a TGH specific hospital capacity forecast and to inform leadership that TGH would have enough capacity to handle the surge. As other health systems were canceling operations, based on information the models presented, TGH leadership decided to continue surgical procedures allowing for continuity in patient care and financial stability of the health system. With the data monitored 2-3x a week, TGH was able to strategically allocate resources, supplies, and staff to accommodate for the surge. The model was directionally reliable enough to generate and execute on this strategic COVID capacity plan. Note, the forecasts made in late July should not be taken as predictive of the likelihood future epidemic waves will or will not exceeding ICU capacity. The outcome of this work demonstrates that epidemiological forecasting, when performed at the county level can have a significant impact on the strategic planning of a COVID response for a county and a hospital.

## Data Availability

All data is available through USA Facts. TGH data is available at link below.

https://www.hillsboroughcounty.org/en/residents/public-safety/emergency-management/stay-safe/covid-19-dashboard

https://usafacts.org/visualizations/coronavirus-covid-19-spread-map

## Acknowledgements

The authors would like to acknowledge valued discussions with Kun Hu and Stefan Edlund.

## Author contribution

JHK and AD conceived the work, JHK, VG, and SP created the model, VG and SP created the automation pipeline, VG, SP, RS, JHK, SB, PH and AD, analyzed the USAFacts COVID data, JHK, SB, VG, SP and SK performed the COVID model analysis, EH, MD, PH and SK analyzed the ICU data, EH, MD, and PH performed the ICU forecast, all authors wrote the manuscript.

## Supplement

### Supplemental Methods

#### Supplemental Data Description

USAFacts COVID-19 datasets^9^ were used for model optimization. The USAFacts dataset is widely used for modeling and analyses by many researchers^10,11,12^ and is also used in aforementioned CDC challenge^7^. In addition to the daily cumulative cases of incidences and death at county level, it also records the number of cases that are currently “unassigned” to any county in a state, thus, enabling one to roll up the numbers to state level for modeling.

#### Hospitalization and Intensive Care Unit Forecasting Model

To support regional collaboration, COVID-19 hospital and ICU admissions were computed from daily predicted incidence as a post process using rate equations determined from local hospital data. Hospital data and models do not represent any patient specific information (no PHI). All data sets used are publicly available through Florida Public Health via an open API that provides aggregated data at the Hillsborough county level^1^. Rate parameters were calculated and fitted using the local hospital data and observations related to hospitalized patients with COVID-19. Rate equations were updated and re-fitted approximately weekly, and more frequently during the second incidence peak in July. Over each independent projection of incidence, the average rate (weighted by incidence) of hospitalization per case (incidence) was 0.35±0.04 and the weighted average rate of ICU admission per hospitalized patient as 0.10±0.05. The rates for each projection period are available in the supplemental data.

#### Forming County Clusters

A major concern for modeling at fine grained resolution is possible sparsity of data. For example, cases from one county may be may be aggregated with and reported in a neighboring county. One solution is to cluster neighboring regions and to aggregate data at the fine grained granularity. However, care should be taken that the process of amalgamating these finer resolution areas does not dilute the dynamics of the epidemic and/or violate the assumption of a well mixed population.

Towards this end, one could use statistical areas like Metropolitan Statistical Area (MSA) as defined by U.S. Census statistics or like Combined Statistical Area (CSA) and Core-Based Statistical Area (CBSA) as defined by Office of Management and Budget. However, the challenge in using these county groupings is that such statistical area spans multiple states. Furthermore, these statistical areas do not cover all the counties in the U.S.

Consequently, we need to develop a custom mechanism to aggregate the counties in such a manner that it:

- Preserves the population mixing characteristics,
- Is sensitive to state boundaries,
- Dynamically identifies the number of correct clusters/groupings of the counties in each state.

A deeper perusal of these above requirements highlights two important facets in formulating this problem - (i) A need to capture the people-people interaction between counties, and (ii) Whether there exist an inter-county affinity or whether the population mixing is primarily intra-county. With regard to the second point, an example of the former (i.e., inter-county affinity) would be suburban regions and satellite township centred around large or economically major cities while for the latter (i.e., inter-county affinity) it would be large rural counties which usually tend to be self contained. It is worthwhile to reiterate here that although this definition is very similar to that of the CBSA, CBSA does not respect state boundaries nor covers all the regions in the U.S.

Hence, we use the census bureau’s 2011-2015 5-Year ACS commuting flows between residence county to workplace county^2^. This data is transformed into a weighted graph (with self-loops), where the weights are the commuter flows. The resulting asymmetric graph is then symmetrized. We use Louvain modularity algorithm^24^ that extracts communities using a greedy optimization approach. The algorithm first detects smaller communities and then progressively aggregates them if the resultant modularity value of the agglomeration is more than that of the individuals. This allows us to avoid specifying the number of communities/clusters to be extracted in advance. In other words, the groups of counties are derived purely based on the work-site to residence mobility patterns.

This approach results in a total of 685 groups of counties which we call county clusters.

#### Model Definition

The compartments shown in figure 1 are listed in Table 2. The flow diagram of figure 1 translates into a system of ODE’s reported in Eqns. 2.

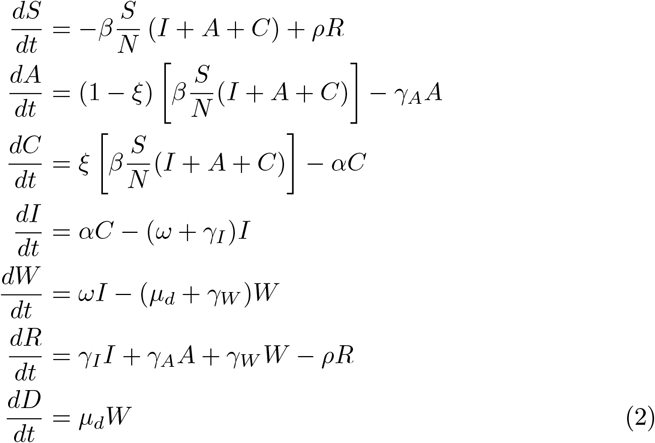

**Table 2:** Table of epidemiological compartments for the model in Figure 1.

| Compartment | Description | Initialization |
| --- | --- | --- |
| S | Susceptible | grid search |
| A | Asymptomatic infectious | algorithmic |
| C | Pre-symptomatic infectious | algorithmic |
| I | Infectious (clinical) | grid search |
| R | Recovered | grid search |
| W | Worsening Symptoms (not infectious) | 0 |
| D | Disease Death | 0 |

An important variable in the SACIR model is the case reporting rate, *ξ*. This parameter partitions new incidence across two paths; symptomatic infectious and asymptomatic infectious. The name suggests that asymptomatic cases are not reported whereas symptomatic cases are. However, the true fraction of reported cases is affected by a convolution of factors (including, but not only, the rate of asymptomatic infection). These includes cases that were truly asymptomatic, cases with symptoms so mild as to not result in an clinical encounter, and/or the efficiency of reporting process itself. For seasonal influenza the fraction of cases registered by public health has been reported to be as low as 3% in some jurisdictions^29^. The asymptomatic fraction of SARS-CoV-2 has been reported to be in a very wide range of 15-90%^52,55,27,43^. The reporting parameter is also affected by the underlying assumption that the population in a region being modeled is well mixed. If one constructs a model where a very large region, hence where the density of people is low, is treated as one well mixed node, this is a source of prediction error. Consider, for example, a patch model where a large country (Admin level 0) is treated as one well mixed region. The model puts the entire population of that country in one patch. But, if the actual epidemic is concentrated in one sub-region or city, that scenario has a built in error. Since the entire population is not well mixed, the denominator in the mass action terms, that is, the parameter *N* in Eqs. 2, should be the population of the affected province or city - not the whole country. In treating reference data from a single city as if it represents an entire country, simplex solver will compensate by decreasing the case reporting rate to an unrealistically low value. Therefore, derived parameter values can be influenced by artifacts related to spatial resolution. The *A* compartment (see figure 1) includes all non-reported incidence.

The symptom appearance rate (see Table 4) contributes a latency between infection and reporting (similar to incubation rate in an SEIR model) with pre-symptomatic individuals also contributing to the force of infection. Case report and mortality data indicate a delay between the infectious period and reported disease mortality. The base SACIR model does not capture this additional latency. Therefore another compartment (W) was added between (I) and (D), as shown in figure 1. This compartment, along with the corresponding “worsening rate”, *ω*, provides for that latency. Moreover, it also describes the fraction of patients whose symptoms become more severe after the clinical infectious period, and is a proxy for demand of clinical intensive care units (ICUs). Individuals in this compartment do not contribute the force of infection, as they are assumed to be clinically isolated in an ICU. Background birth and death rates are assumed to be zero. The differential equations for the SACIWR model are shown below. Model integration was performed with an order 8(5,3) Dormand-Prince solver from the Apache software foundation, and implemented in the STEM framework^33^.

#### Model Parameters and Parameter Estimation

Epidemiological parameters were obtained by a combination of Nelder-Mead simplex, grid search, and grid search around literature estimates as shown in Table 4.

As *R*_0_ is a ratio of transmission rate to recovery rate(s), (please refer to Equation 1), we need to vary only one of these two parameters to avoid over fitting. We chose to vary the transmission rate and set the recovery rate(s) based on literature estimates. Please also recall that in our approach, the transmission rate is time sensitive in the sense that we have multiple transmission rate to account for the different interventions. However, the recovery rate(s) are the same for the entire time period, as it is the characteristics of the disease and there has not been a significant change in the treatment regimen. A recent paper^40^ has estimated that the recovery time of the asymptomatic individuals is roughly the same as that of those who exhibit symptoms, which is about 14-20 days. However, in our case, the infected individuals are split into two categories, compartment *I* and compartment *W*, where the latter represents patients with advanced stage of illness. Consequently, we split the recovery rates into three, asymptomatic recovery rate (*γ*_*a*_), recovery rate for *I* (*γ*_*i*_) and, recovery rate for *W* (*γ*_*w*_). *γ*_*a*_ was set based to be 16 days, close to the midpoint region of the range provide in^40^. *γ*_*i*_ was set to be on the lower extreme of the range - 14 days, and *γ*_*w*_ was set to the upper limit of the range - 20 days^45^. It is important to highlight that since we vary the transmission rate over a range of continuous values, and as there are multiple transmission rates to fit the different portions of the data, the impact of the choice of recovery rates is mitigated so long as it in the clinically valid range.

The transmission rates were allowed to vary from 0.02 to 0.85, which we determined through experiments covered all the ranges possible based on the data we had at the time of the experiment. Immunity loss rate was fixed based on an estimated 10 months period of infection^38^.

As mentioned earlier, Nelder-Mead optimization algorithm was used to fit the variable parameters and the algorithm was run multiple times with reinitializations until a convergence in the parameter values was obtained across two runs. This was done to prevent trapping at local minima and for the reproducibility of the parameter values; thus, ensuring that analysis of these parameters across regions are not confounded by numerical variations of the algorithm and reflect actual variation in epidemic dynamics. The average fitted nrmse at each resolution is also listed in Table 4. Figure 2a measures the distribution of each of the optimized parameters across regions through a combination violin-plot, box-plot. The distribution for all other optimized parameters is shown in the supplement figure S1. L-statistics obtained from the box plot are listed in Table 4 for each parameter for comparing the distributions across spatial scales. The majority of the epidemiological parameters shown in figure 1 are variable parameters with values determined by the AI optimization pipeline. The parameters were fitted using the data starting from the first day of the reported infection up to August 3rd, 2020. The evaluation MAPE was measured with respect to reference data reported for the following 4 weeks from August 3rd, 2020. Parameter values with uncertainties or range are provided in the Results. All parameter values were obtained independently for all regions at three spatial resolutions (county, county cluster, and state). Resulting *R*_0_ values are then compared to recent estimates from the literature (see Table 5). Over the course of the epidemic, changes in social distancing policies and other non-pharmaceutical interventions (npi) are manifest by a reduction in transmission rate or changes in other epidemiological parameters. These modified values reflect the time varying effective reproductive number, *R*_*eff*_ (or *R*_*t*_). *R*_*eff*_ changes with time due to depletion of the susceptible population as well as widespread policies (and policy removals). The maximum *R*_0_ measured early in the epidemic reflects the *“natural”* infection dynamics give *“normal”* social interactions, absent npi.

#### Hyper-local Pipeline

To provide a scale-able architecture around the STEM model described in the sub-section Base Model, we built an automation pipeline that performs a variety of task ranging from data ingestion, smoothing to invoking the model and post-processing (as required).

The pipeline begins with a data set interface that allows the users to connect to multiple data set providers and convert them into a unified representation for the various downstream processes. For this paper, we used the USAFacts data set interface. The downloaded data is then pre-processed to fix any data quality issues like negative daily cases and data smoothing. The pipeline provides access to different smoothing algorithms like Moving Average, Savitzky Golay and also Adaptive Savitzky Golay filters. For the purpose of this paper, we use Adaptive Savitzky Golay filter as it allows us to determine the right polynomial for smoothing with minimal data loss.

It is to be noted that as the pandemic evolves, there are number of responses by governments and individuals at various levels. In the case of COVID-19, such responses have mainly been through Non-Pharmaceutical Interventions (NPIs), which mainly aim at promoting social distancing and reducing the transmission risks. Further, these NPIs have different level of enforcement and consequently its effects are manifested as various discontinuities on the case incidence and death curves. Another source of discontinuities are due changes in mobility. These discontinuities in the curve can be thought to be different stages of the pandemic and warrants the model parameters to be sensitive to it. We use these discontinuities to define “segments”^48^ and use the corresponding start and end times as a part of Nelder-Mead optimization of transmission rate versus time. The pipeline also supports multi-processing, where each region is run by a separate instance of a docker image; thus, allowing easy scaling of the clusters based on the requirement.

#### Models evaluated for comparison

Our selection criteria for comparing to the models participated in the CDC COVID-19 forecasting challenge^4^ was based on the availability of the models’ forecast submission on August 3^rd^, 2020. Only the models that had submission for both deaths and cases were included in our comparison regardless of the type of the model. The other model selection criterion was providing the forecasts (both deaths and cases) for up to four weeks ahead. Additionally, for the state-level comparison, only the models that provided forecast for all the 50 states (excluding the District of Columbia) were included. List of the models with forecasting submission to the CDC challenge on August 3^rd^, 2020, that either met or did not meet the aforementioned criteria, can be found in Table 3.

**Table 3:** CDC Challenge Models (submitted 8/3/20). Data types C=cases, D = Deaths. ⊆ indicates subset. Models used for comparison provided predictions for both data types and all regions.

| Model Name | Methods | data types | included ? | missing data |
| --- | --- | --- | --- | --- |
| Auquan<sup>3</sup> | Fitted SEIR | D | No | C |
| CDDEP<sup>5</sup> | Bayesian SEIR | C | No | D |
| CMU<sup>6</sup> | Autoreg. time-series | D | No | C |
| Columbia<sup>7</sup> | Metapop SEIR | C,D | No | $\subseteq$ C |
| Columbia-UNC<sup>57</sup> | Stat. survival-conv. | D | No | C |
| Covid19Sim<sup>8</sup> | SEIR | C,D | Yes | n/a |
| DDS<sup>9</sup> | Bayesian hierarchical | C,D | Yes | n/a |
| ERDC<sup>10</sup> | SEIR | C,D | Yes | n/a |
| ESG<sup>11</sup> | Fitted gaussian dist. | D | No | C |
| Geneva<sup>12</sup> | Statistical model | C,D | No | $\subseteq$ weeks |
| GT-CHHS<sup>37</sup> | Agent-based | D | No | C |
| GT-DeepCOVID<sup>47</sup> | Deep learning | D | No | C |
| IHME<sup>13</sup> | regression model | C,D | Yes | n/a |
| ISU<sup>14</sup> | Spatiotemporal model | C,D | No | $\subseteq$ regions |
| JCB<sup>25</sup> | Statistical model | D | No | C |
| JHU<sup>15</sup> | Metapop. SEIR | C,D | Yes | n/a |
| Karlen<sup>36</sup> | Discrete time | C,D | Yes | n/a |
| LANL<sup>16</sup> | Statistical model | C,D | Yes | n/a |
| LNQ<sup>58</sup> | Machine learning | C | No | D $\subseteq$ weeks |
| LSHTM<sup>21</sup> | Ensemble (3 models) | D | No | C |
| MIT-CovAlliance<sup>22</sup> | SIR | C,D | No | $\subseteq$ regions |
| MIT-ORC<sup>41</sup> | SEIR | C,D | Yes | n/a |
| MOBS<sup>56</sup> | Metapop. SLIR | D | No | C |
| NotreDame-FRED<sup>30</sup> | Agent-based | D | No | C |
| NotreDame-Mob<sup>31</sup> | SEIR | D | No | C |
| Oliver Wyman<sup>39</sup> | SIR | C,D | No | $\subseteq$ weeks |
| PSI<sup>53</sup> | Stoch SEIRX | D | No | C |
| QJHong<sup>34</sup> | ML | D | No | C |
| RPI-UW<sup>50</sup> | SIR | D | No | C |
| STH<sup>35</sup> | Stat growth | D | No | C |
| UA<sup>23</sup> | SIR | D | No | C |
| UCLA<sup>61</sup> | mod SEIR | C,D | Yes | n/a |
| UCM<sup>17</sup> | SEIR | D | No | C |
| UM<sup>18</sup> | Ridge regression | C,D | No | $\subseteq$ regions |
| UMass-MB<sup>19</sup> | Mechanistic Bayesian | C,D | No | $\subseteq$ regions |
| USC<sup>51</sup> | SIR | C,D | Yes | n/a |
| UT<sup>59</sup> | Bayesian | D | No | C |
| UVA<sup>20</sup> | ensemble (3 models) | C | No | D |
| YYG<sup>32</sup> | SEIR and ML | D | No | C |
| Ensemble | ensemble | C,D | No | included above |

### Supplemental Results

#### Derived Epidemiological Parameters

As discussed in the main manuscript, derived recovery rates (Table 4) are within the range of current clinical studies.^20,21,22,23,24^ These studies document pre-symptomatic infections 5-14 days from the time of exposure.^20,21,22,23,24^ After symptom onset, recovery of replication-competent virus has been documented 10 − 20 days after symptom onset corresponding to a recovery rate between 0.05 − 0.1 [days^−1^]^54^. Another study documented shedding of virus up to 8 days following the symptomatic period^26^.

**Table 4:** Table of epidemiological parameters. NM = Nelder-Mead. GS=Grid Search.

| Parameter | L-stat. | States | County<br>Clusters | Counties | method<br>√source |
| --- | --- | --- | --- | --- | --- |
| trans<br>rate<br>(max $\beta$) | q3<br>median<br>mean<br>q1 | 0.341<br>0.274<br>0.293<br>0.226 | 0.348<br>0.236<br>0.298<br>0.171 | 0.231<br>0.111<br>0.191<br>0.086 | NM |
| trans<br>rate<br>(all $\beta$) | q3<br>median<br>mean<br>q1 | 0.177<br>0.120<br>0.145<br>0.082 | 0.192<br>0.120<br>0.155<br>0.065 | 0.173<br>0.111<br>0.145<br>0.072 | NM |
| case<br>reporting<br>rate<br>$\xi$ | q3<br>median<br>mean<br>q1 | 0.061<br>0.053<br>0.050<br>0.042 | 0.060<br>0.050<br>0.048<br>0.021 | 0.061<br>0.050<br>0.049<br>0.017 | NM |
| symptom<br>appear.<br>rate<br>$\alpha$ | q3<br>median<br>mean<br>q1 | 0.109<br>0.098<br>0.101<br>0.093 | 0.108<br>0.098<br>0.097<br>0.079 | 0.108<br>0.098<br>0.097<br>0.065 | NM |
| worsening<br>rate<br>$\omega$ | q3<br>median<br>mean<br>q1 | 0.161<br>0.104<br>0.145<br>0.067 | 0.197<br>0.097<br>0.179<br>0.041 | 0.212<br>0.075<br>0.185<br>0.017 | NM |
| infectious<br>mortality<br>rate<br>$\ddagger \mu_d$ | q3<br>median<br>mean<br>q1 | 0.009<br>0.004<br>0.006<br>0.002 | 0.008<br>0.003<br>0.006<br>0.001 | 0.008<br>0.003<br>0.007<br>0.001 | NM |
| recov $\gamma_A$<br>recov $\gamma_I$<br>recov $\gamma_W$ | *fixed*<br>*fixed*<br>*fixed* | 0.06<br>0.071<br>0.05 | 0.06<br>0.071<br>0.05 | 0.06<br>0.071<br>0.05 | *lit.*<sup>45</sup><br>GS<br>0.048-0.071 |
| imm.<br>loss<br>$\rho$ | *fixed* | 0.1<br>$mos^{-1}$ | 0.1<br>$mos^{-1}$ | 0.1<br>$mos^{-1}$ | $(8-12)^{-1}$<br>$mos^{-1}$<br>*lit*<sup>38,46</sup> |
| $\langle nrmse \rangle$ | mean | 0.038<br>$\pm 0.008$ | 0.065<br>$\pm 0.040$ | 0.093<br>$\pm 0.052$ | NM |

<sup>†</sup> rate units are per day unless otherwise indicated.
<sup>‡</sup> disease death rate (per day) not to be confused with case fatality rate (cfr)

**Figure S1:**
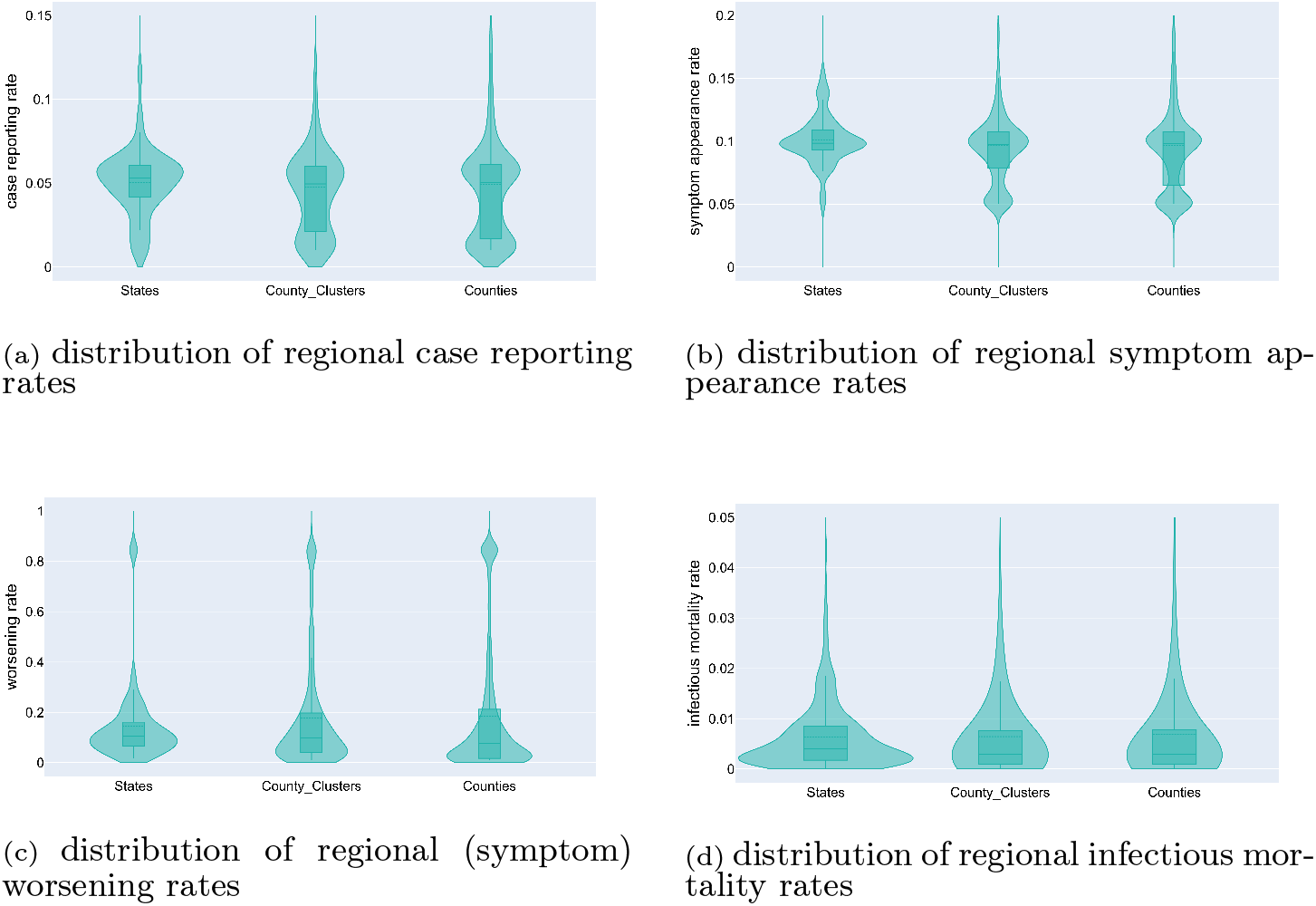
Distribution of regional values of parameters obtained via Nelder-Mead Simplex model optimization. The distribution of transmission rates and derived reproduction numbers are shown in manuscript figure 2a and 2b. Parameters vary by region and time as the disease dynamics change locally in response to non-pharmaceutical interventions and other local factors.

Table 5 provides statistics from the measured distribution of reproduction number values obtained using the epidemiological parameters in Table 4 at each spatial resolution. From the distributions shown in figure 2b the Table lists four L-statistics; first quartile, mean value, median value, and third quartile of the measured distributions for *R*_0_ and *R*_*eff*_ as defined above. The Table also shows other estimates of the reproduction number from the literature. The examples from the literature are simply ordered based on the upper limit for each reported range (and not related to the L-statistic categories). The median value for *R*_0_ systematically decreases as the spatial resolution of the model increases from state to county. The distributions labeled *R*_*eff*_ (which reflect the diverse range of non-pharmaceutical interventions and range of compliance across all regions and time) do not show a systematic shift.

**Table 5:** This table shows the derived values of the maximal basic reproductive number of Sars-CoV-2, by spatial resolution, computed from the epidemiological parameter values in table 4. The final column shows estimates from current literature.

| Param | L-stat. | States | County<br>Clusters | Counties | *Lit* <sup>†</sup> |
| --- | --- | --- | --- | --- | --- |
| $R_0$ | $q_3$ | 7.216 | 7.929 | 5.256 | *up to 9.24*<sup>60</sup> |
| from | median | 6.388 | 5.530 | 2.909 | *1.3 - 6.5*<sup>49</sup> |
| max | mean | 6.528 | 6.770 | 4.442 | *2.4 - 3.1*<sup>28</sup> |
| $\beta$ | $q_1$ | 4.966 | 3.688 | 1.790 | *1.6 - 3.0*<sup>44</sup> |
|  |  |  |  |  | *2.03 - 2.77*<sup>42</sup> |
| $R_{eff}$ | $q_3$ | 3.942 | 4.260 | 3.975 | |
| from | median | 2.610 | 2.642 | 2.550 |  |
| all | mean | 3.222 | 3.511 | 3.371 |  |
| $\beta$ | $q_1$ | 1.833 | 1.436 | 1.602 | |

<sup>†</sup> Examples from literature are ordered by reported upper limit

#### Hospitalization and Intensive Care Unit Forecasting Results

Manuscript figure 6 shows predicted ICU demand for Tampa General Hospital (TGH), Hillsborough County, FL. Figure S2a shows the regular incidence predictions for the county (from the SACIR model), and figure S2b shows the predicted COVID-19 hospitalizations. The green curve is actual data and the magenta curve (and solid points) show independent projections made at least weakly beginning in April 2020.

**Figure S2:**
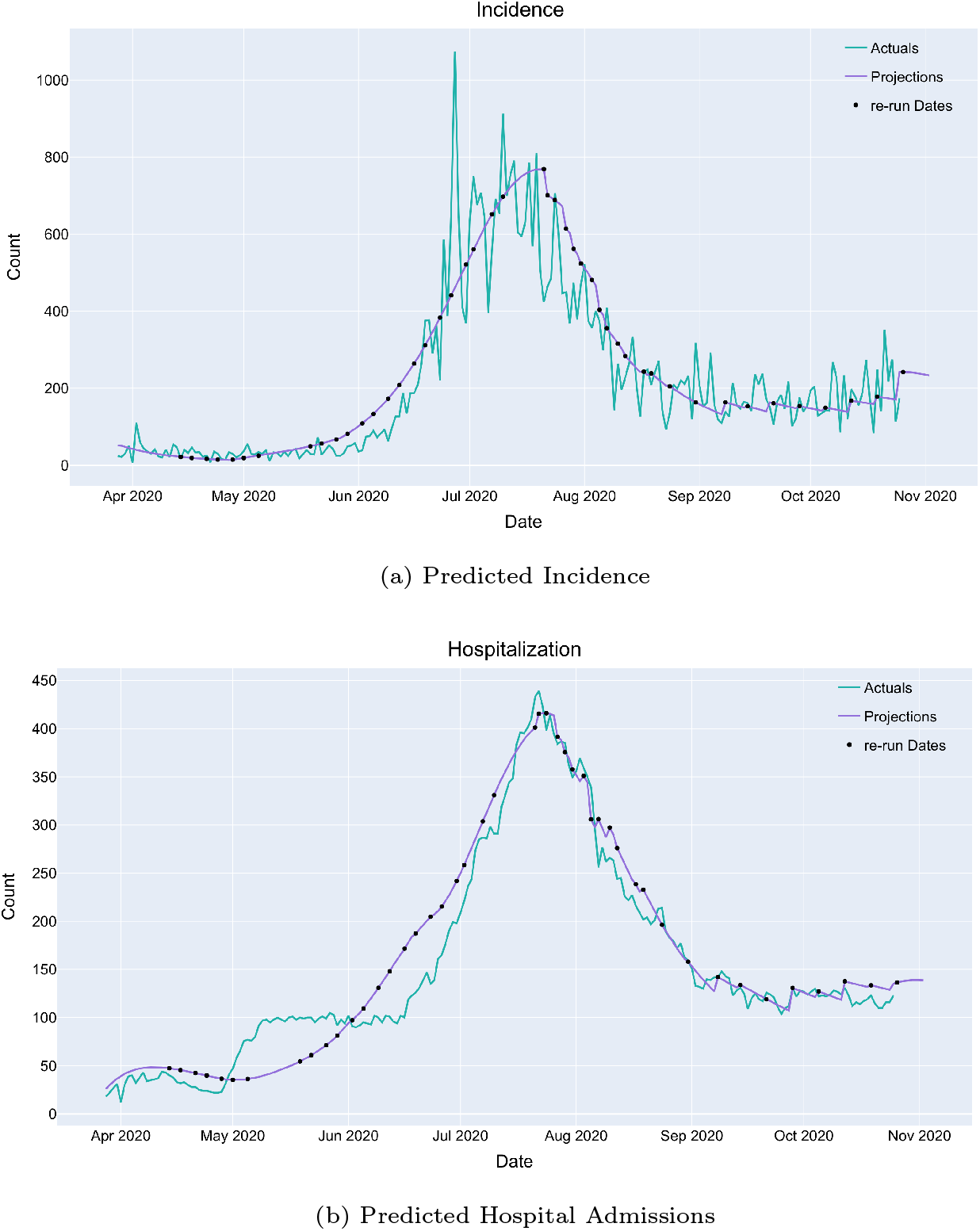
Projected incidence and projected hospitalizations for Tampa General Hospital in Hillsborough County, Florida, USA.

